# The Role of Type 2 Diabetes in Shaping Multimorbidity Progression: Evidence from the UK Biobank Cohort

**DOI:** 10.64898/2026.05.01.26352200

**Authors:** Jie Zhang, Lasse Bjerg, Susanne Boel Graversen, Henrik Støvring, Christina C. Dahm, Bendix Carstensen, Daniel Witte

## Abstract

**Aims/hypothesis:** Type 2 diabetes (T2D) frequently co-occurs with other chronic conditions and has been suggested as a key driver of multimorbidity development. However, the temporal dynamics of how T2D influences multimorbidity progression are not well understood. We aimed to investigate how T2D influences the rate of multimorbidity progression.

**Methods:** We analyzed data from the UK Biobank, a prospective population-based cohort study (n=502,368, median age 58 years [range 37–73 years], 46% male at baseline) with a median follow-up of 15.3 year. 8.7% of participants were diagnosed with T2D over the follow-up period. We counted the current number of morbidities (among 80 long-term chronic conditions) identified through hospital admission records using ICD-10 diagnosis codes. We modeled the age-specific transition rates between multimorbidity states using multistate models tailored for time-split data (i.e., 2 to 3 morbidities, T2D (with 1 morbidity) toT2D (with 2 morbidities), etc.). Age was modeled using natural splines with an interaction term with T2D, adjusting for sex, education, ethnicity, smoking, and body mass index.

**Results:** The total follow-up time was 7.5 million person-years (PY), of which 0.33 million PY was in T2D. Individuals with T2D consistently experienced higher transition rates between morbidity transition states. For example, the transition rate from 2 to 3 morbidities was 7.94 per 100 PY without the presence of T2D, compared to 18.35 per 100 PY with T2D (rate ratio=2.31, 95% CI: 2.28-2.34). The excess in transition rates was most pronounced from states with few comorbidities. Further, the transition rates were consistently influenced by T2D status and age, with younger individuals with T2D showing the most accelerated progression.

**Conclusion/interpretation:** Our study suggests that T2D is associated with accelerated development of subsequent multimorbidity, highlighting T2D as a critical contributor of chronic disease accumulation. The progression of disease accumulation is more pronounced in younger age groups, which suggests different underlying mechanisms of multimorbidity progression across age, warranting further investigation. The findings indicate the need for early intervention among younger people with T2D to slow multimorbidity progression.

## Introduction

People with type 2 diabetes mellitus (T2D) face a significantly higher risk of developing multiple long-term conditions (MLTCs) compared to the general population[1], [2]. Studies report that up to 52% and 98% of people with T2D have at least one other chronic condition, depending on population characteristics and time since diagnosis[3]. In addition to micro- and macrovascular complications that reflect shared underlying pathophysiology with T2D, T2D is also associated with a wider range of discordant conditions across multiple organ systems. These conditions include cancer, chronic obstructive pulmonary disease (COPD), non-alcoholic fatty liver disease, dementia, and other mental health diseases[4], [5]. Several biological mechanisms may explain the link between T2D and MLTCs. Prolonged exposure to elevated glucose and insulin resistance impairs the vascular and nervous systems, promoting systemic inflammation and oxidative stress[6]. T2D is also linked to accelerated biological aging, characterized by a reduced physiological resilience, increased susceptibility to disease, and progressive multi-organ dysfunction[7]. Together, these pathological processes may activate or amplify disease pathways across organ systems, thereby facilitating the accumulation of comorbid chronic conditions over time in people with T2D.

The development of T2D-related MLTCs is a dynamic process, with T2D potentially arising at any point along the trajectory of MLTC accumulation. For instance, T2D may develop before CVD, accelerating atherosclerosis and increasing vascular risk, or it may emerge after a cardiovascular event, as a consequence of post-event metabolic dysregulation, inflammation, and reduced physical activity. This bidirectionality means that the temporal ordering of conditions matters clinically. Yet most existing studies treat T2D as a static exposure and only adjust for the number of comorbidities at baseline, which limits the ability to characterize the dynamic pathway through which multimorbidity accumulates over time.

Multistate modelling offers a framework to address these complexities. A key advantage is the ability to explicitly capture transitions between multiple disease states as they unfold over time, while accounting for concurrent complication burden[8], [9]. Specifically, multistate models can quantify transition rates and the probability of occupying a given state at specified time points. This approach has been applied to examine progression in diabetes-related complications across clinical profiles[10], [11], [12].

In the current study, we extended this approach to examine the dynamic influence of T2D on the accumulation rate of 80 long term conditions (LTCs). The method accommodates T2D as a time-varying exposure, allowing its association with LTC accumulation to vary according to current disease burden and age[13]. Unlike previous multistate studies restricted complications sharing common pathophysiological mechanisms with T2D, our analysis encompassed a broad range of 80 LTCs, capturing both concordant and discordant conditions.

## Methods

### Study design and population

UK Biobank is a population-based health research resource with approximately 500,000 people, aged between 37 years and 73 years at the time of entry between 2004 and 2010 across the UK. A full description of the study design, participants, and quality control methods has been described in detail previously[14], [15]. The study is reported in accordance with the Strengthening the Reporting of Observational Studies in Epidemiology (STROBE) guidelines[16].

### Ethics approval

UK Biobank received ethical approval from the Research Ethics Committee (REC reference for UK Biobank is 11/NW/0382). All participants gave written informed consent before enrolment in the study, which was conducted in accordance with the principles of the Declaration of Helsinki. Secondary analysis of UK Biobank data does not require additional ethical approval, as it is covered under the original UK Biobank ethics framework. This study has been conducted using the UK Biobank Resource under Application Number 81520.

### T2D and MLTC

T2D diagnosis was defined as at least one recorded ICD-10 code for T2D (E11) in available Hospital Episode Statistics (HES) data. In addition to T2D, we identified the presence of 79 other LTCs using ICD-10 diagnosis codes and OPCS-4 procedure codes from HES[17], [18]. Consistent with our previous work[19], LTCs were grouped into the following disease categories: blood and blood-forming organs; circulatory system; digestive system; endocrine, nutritional and metabolic diseases; genitourinary system; infectious and parasitic diseases; mental health; musculoskeletal system; neoplasms; nervous system; respiratory system; and symptoms and signs. Full details of all conditions and their corresponding codes are provided in Table S1 and https://github.com/arborzhang/ukb-tracking/tree/main/MLTC_code.

### Sociodemographic characteristics and lifestyle factors

Sociodemographic characteristics included age at baseline, sex (male/female), ethnicity (White/Black/Asian/other), and education level (college or above; A levels; O levels, GCSEs or equivalent; NVQ, HND, HNC, or equivalent; other professional qualifications; or none of the above (equivalent to less than high school diploma and CSEs or equivalent)). Smoking status was self-reported at baseline and classified as current, former, or never smoker. BMI was calculated as weight in kilograms divided by height in metres squared and categorised according to WHO recommendations as normal weight (<24.9 kg/m^2^), overweight (25.0–29.9 kg/m^2^), and obesity (≥30 kg/m^2^). All sociodemographic and lifestyle variables were ascertained through self-completed questionnaires administered at baseline.

### Statistical analysis

Baseline characteristics were summarized across three mutually exclusive groups: those with T2D, those without T2D but with at least one other LTC, and those without any LTC. Continuous variables were summarized using means (standard deviation [SD]) or medians (interquartile range [IQR]), and categorical variables using counts and percentages.

### T2D and LTC accumulation rate

For each participant, we constructed longitudinal records including birth date, cohort entry and exit dates, the first recorded diagnosis date of each LTC. This allowed us to track the chronological sequence of disease accumulation over time. LTC accumulation was modelled as progression through states defined as number of LTC(0 to 8+ LTCs) or death. The type of LTC was not discerned, only the cumulative count of LTCs was considered. All participants were followed from enrolment until death, loss to follow-up, or end of study (31 December 2023), whichever occurred first.

Using a Lexis based approach we split person-time at each LTC diagnosis date[20], allowing participants to transition between successive LTC states and, when applicable, transition from the non-T2D to the T2D pathway upon diagnosis. A multistate model was used to estimate transition rates between successive LTC states[21]. The model comprised 19 states defined by LTC count (0→8+ LTCs), cross classified by T2D status, plus death as an absorbing state (Figure 1). Participants without T2D could progress to the next LTC state (e.g., 1→2 LTCs) or develop T2D (e.g., 1LTC→T2D+1LTC); once in the T2D pathway, persons could only progress forward through T2D-specific states (e.g., T2D+1LTC→T2D+2 LTCs) or die. Death was treated as an absorbing state accessible from any state throughout follow-up. To account for temporal variation in transition rates, the dataset was further subdivided by single calendar year. Transition intensities for all possible transitions were estimated simultaneously using maximum likelihood estimation within the multistate framework. The full transition matrix is presented in Table S2.

**Figure 1.**
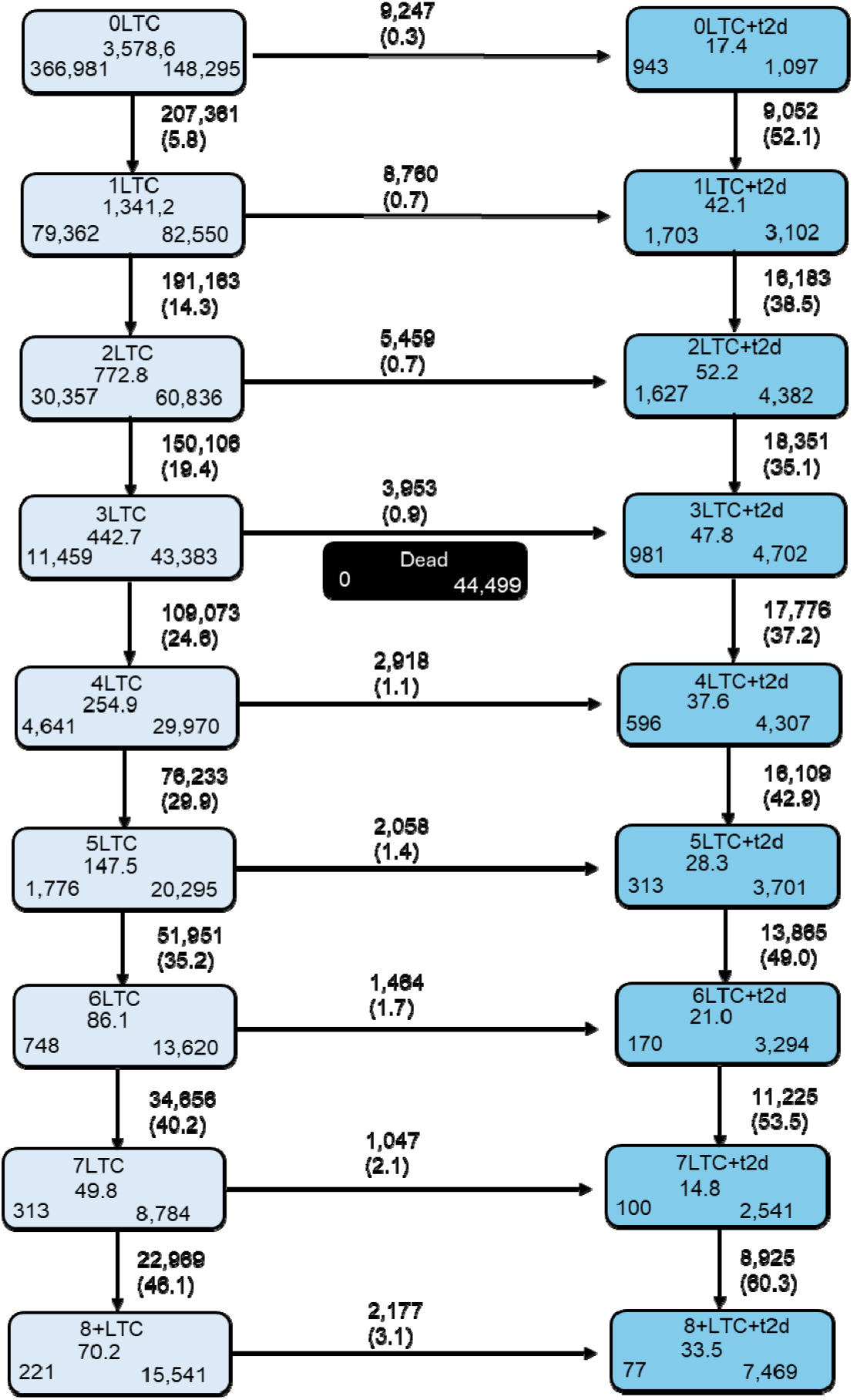
MLTCs progression states with and without T2D: A multistate Markov model. The model assumes a progressive process in which individuals can only gain a single LTC at a time and cannot revert to a previous state, with death as the absorbing (final) state. Numbers at the bottom of each box denote the number of people who begin, respectively end follow-up in that state. Transitions between states are indicated by arrows. Numbers by arrows are the number of transitions and transition rates per 1000 person-years of follow-up. n=502,368 individuals contributing 7.5 million person-years of follow-up across all morbidity transitions. *Abbreviations: LTC, long term condition; T2D, type 2 diabetes*

To account for potential diagnostic delays or incomplete baseline ascertainment, we applied a 12-month bandwidth for diagnoses after cohort entry, whereby diagnoses occurring within the first year after cohort entry were considered present at baseline. Incident LTC accumulation therefore began 12 months after cohort entry and participants could enter the cohort with pre-existing LTC diagnoses. Consistent with the assumption of the irreversible nature of chronic diseases, backward transitions (e.g., 2 LTCs → 1 LTC) were not permitted in the multistate framework.

### Model specification

Transition rates were estimated using Poisson models. Three time dimensions were incorporated, including attained age, time since enrollment and calendar time. The effect of attained age on log-rates was modelled using natural splines with knots positioned at empirically derived quantiles based on the distribution of age at transitions. Similarly, the effect of time since enrollment was modeled using natural splines with knots at 1, 5, 10, and 20 years; and the effect of calendar time on log-rates was included as a linear term to account for period effects. The current number of LTCs (T*morb*) was included as a time-varying covariate, with exp(β T*morb*) representing the hazard ratio for acquiring an additional LTC per existing condition.Three Poisson regression models were fitted with log transition rate as the outcome. Model 1 included attained age, sex, time since enrolment, current number of LTCs (T*morb*), a T2D indicator variable, and a T2D × T*morb* interaction term, the latter testing whether the association between T2D and MLTC progression varied by existing disease burden. Model 2 additionally adjusted for ethnicity, education, smoking status, and BMI as potential confounder. Model 3 extended Model 2 by adding an T2D×age interaction term to examine whether the effect of T2D on MLTC accumulation varied with ages. To quantify the T2D-specific association at each level of LTC, we calculated transition rate ratios (TRRs) comparing individuals with and without T2D while holding total LTC count constant. For example, an individual with T2D and one additional LTC (T*morb*=1, total=2) was compared to a participant without T2D but with two LTCs (T*morb*=2, total=2). This approach was applied consistently across all disease states (e.g., T2D+2LTC vs. 3 non-T2D LTCs, T2D+3LTC vs. 4 non-T2D LTCs), thereby isolating the association of T2D with LTC accumulation while accounting for overall disease burden and the competing risk of death.

Mortality was estimated using the same Poisson framework as the transition rate models, with mortality as the outcome of interest.

### Stratification

Analyses were stratified by sex to explore whether the association between T2D and LTC accumulation differed between men and women.

### Sensitivity analysis

In the primary analysis, the number of LTCs was modelled as a continuous variable, assuming a linear relationship between LTC count and transition rates. As a sensitivity analysis, we repeated all models treating the number of LTCs as a categorical variable, allowing each LTC state to have an independent effect without imposing any functional form assumption.

All analyses were conducted in R (version 4.4.0) on the UKB platform. The multi-state models were implemented using the “*Epi*” and “*popEpi*” packages in R.

## Results

The study population comprised 502,368 individuals. During follow-up, 43,616 (8.7%) participants received a diagnosis of T2D. By the end of follow-up, 309,024 (61.5%) had at least one LTC other than T2D, whereas 149,728 (29.8%) remained free of LTCs.

Participants with T2D were older (mean age=59.6 years, SD=7.18), followed by those with non-T2D LTCs (57.9, SD=7.75) and those without any LTCs (52.9, SD=7.83). The T2D group had a higher proportion of men (59%) compared with the non-T2D LTC and no-LTC groups (45% and 43%, respectively). Compared with other groups, participants with T2D had lower educational attainment and household income, higher BMI and waist circumference, and were more likely to be current or former smokers. As expected, HbA1c and fasting glucose levels were markedly higher in the T2D group (Table 1).

After 15.8 years of follow up (IQR=14.6 to 16.1), the most prevalent chronic condition with T2D was hypertension (n=9,127), followed by the triple combination of T2D, hypertension, and CHD (n=4,999). Other common patterns occurring together were T2D with hypertension and osteoarthritis (n=4,258), and T2D with hypertension and gastrointestinal disease (n=3,965) (Figure S1).

### Transition matrix of LTC accumulation

For the non-T2D path, transition rates increased progressively with accumulating conditions, from 14.3 per 1000 PY (1→2 LTCs) to 46.1 per 1000 PY (7→8+ LTCs) (Figure 1). Among individuals with T2D, the transition rate was 52.1 per 1,000 PY at the 1→2 LTC transition, 38.5 and 35.1 per 1,000 PY at the 2→3 and 3→4 transitions respectively, and 53.5 per 1,000 PY at the 7→8+ LTC transition. For individuals without T2D, the rate of developing T2D increased with the number of existing LTCs, rising from 0.3 per 1000 person-years in those with no LTCs to 3.1 per 1000 person-years in those with 8 or more LTCs. A total of 44,499 deaths were recorded during follow-up.

Age at transition increased with the number of LTCs in both groups. In the non-T2D group, median age increased from 66.3 years at the 2→3 transition to 71.7 years at the 6→7 transition; a similar gradient was observed in the T2D group, from 66.4 to 70.7 years across the same transitions. Participants in the T2D group were consistently younger at each transition than their non-T2D counterparts (Table S3).

### T2D and longitudinal LTC accumulation using multistate models

In the multistate models, presence of T2D was consistently associated with higher LTC accumulation in all transition states. For instance, the state-specific TRR was 2.31 (95% CI: 2.28–2.34), at the 2→3 LTC transition, corresponding to transition rates of 7.94 per 100 PY among individuals without T2D and 18.35 per 100 PY among those with T2D. Although absolute transition rates increased with greater LTC burden, the relative effect of T2D attenuated progressively: state-specific TRRs were 1.96 (95% CI: 1.95–1.98) for the 3→4 transition, 1.67 (95% CI: 1.66–1.68) for 4→5, 1.42 (95% CI: 1.41–1.43) for 5→6, 1.21 (95% CI: 1.20–1.21) for 6→7, and 1.02 (95% CI: 1.01–1.03) for 7→8+ LTCs (Table 2, Figure 2).Results remained consistent after adjusting for covariates in Model 2. Male sex, lower education, current smoking and higher BMI were associated with faster LTC accumulation (Table 2, Model 2).

**Figure 2.**
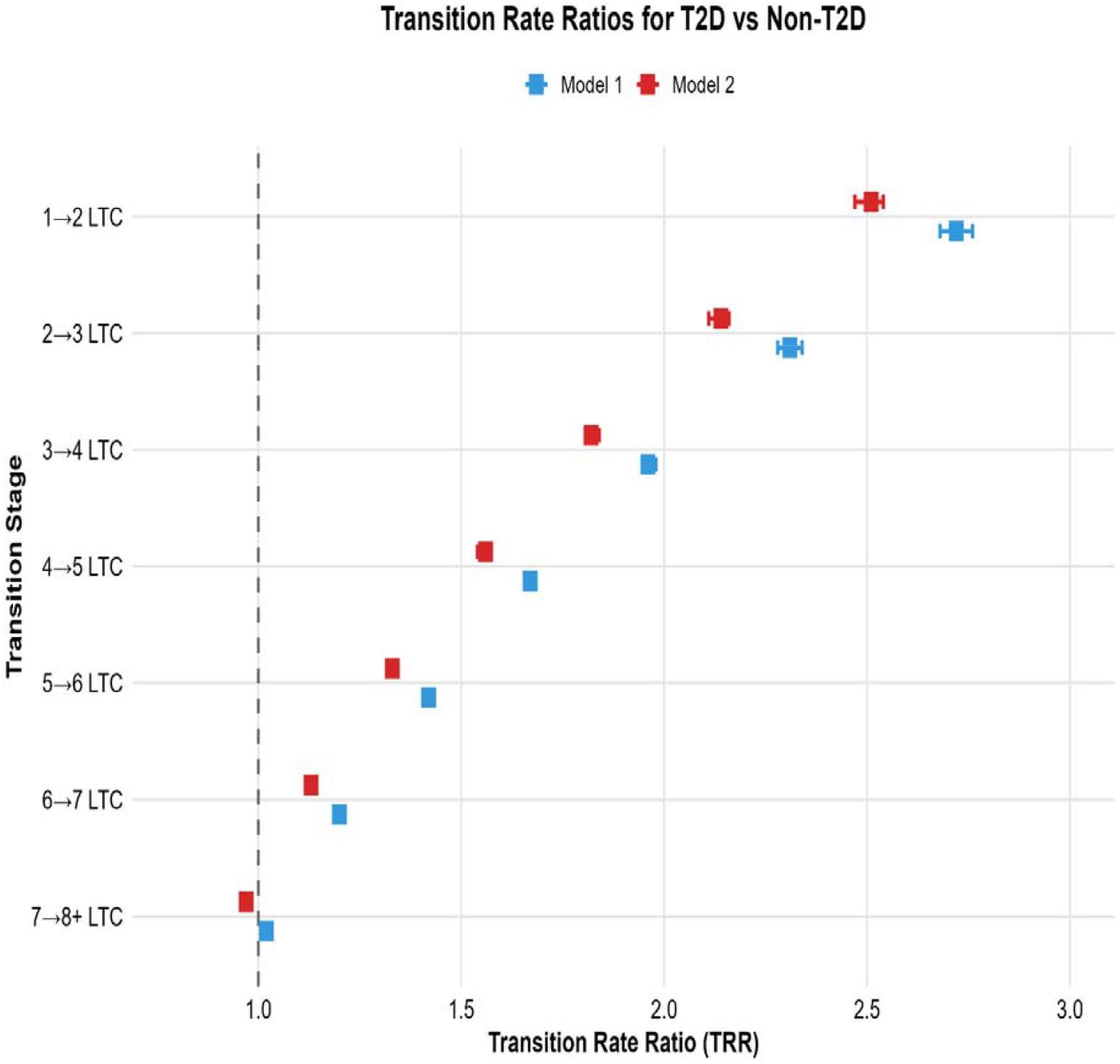
Transition Rate Ratios Comparing Individuals with and without Across Different LTC states. Transition rate ratios (TRRs) for LTC accumulation comparing individuals with T2D to those without T2D. Model 1 was adjusted for age, time since enrolment, calendar time, sex, current number of LTCs, and a T2D × LTC interaction term, allowing the association of T2D with LTC accumulation to vary by disease burden(blue markers). Model 2 further adjusted for ethnicity, education, smoking status, and BMI (red markers). Point estimates are shown as square markers with 95% confidence intervals represented as corresponding coloured bands. *Abbreviations: CI, confidence interval; LTC, long-term conditions; T2D, Type 2 Diabetes; ref, reference*.

Model 3 included an interaction between T2D and age to allow the association varying by ages across different LTC states (Table 3, Model 3). The association between T2D and LTC accumulation was strongest at younger ages and progressively diminished with older age. For transition from 2→3 LTCs, state-specific TRRs were 3.25 (95% CI: 3.16-3.35) at age 50, 2.39 (95% CI: 2.34-2.43) at age 60 and 1.92(95% CI: 1.87-1.95) at age 70, respectively. This age gradient was consistent across all transition states, with TRRs declining at higher LTC states (Figure 3, Table 3). These findings indicate that T2D exerts a stronger accelerating association on LTC accumulation at younger age, with the relative magnitude diminishing with both increasing age and higher LTC burden.

**Figure 3.**
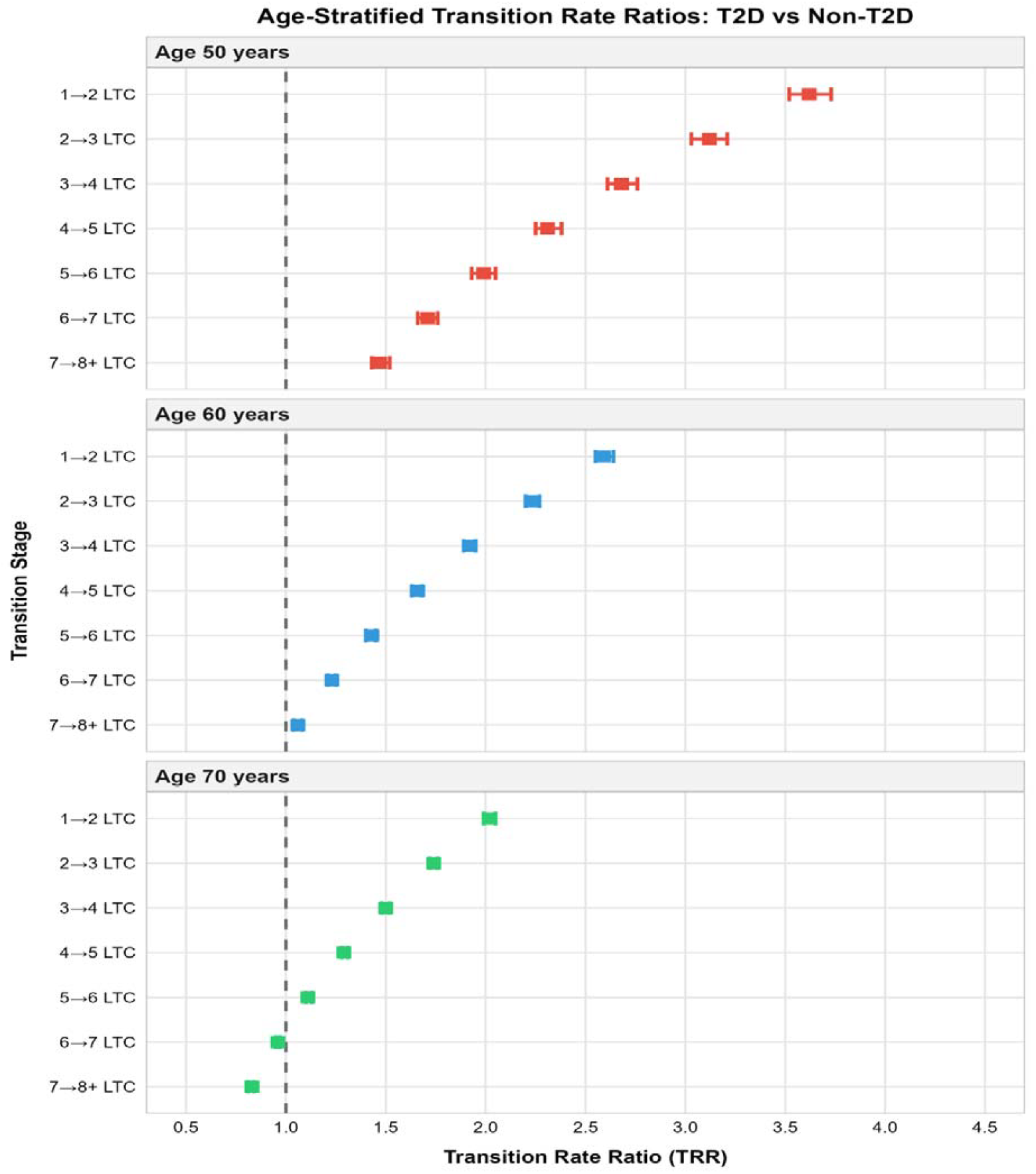
Transition Rate Ratios Comparing Individuals with and without T2D varying by disease burden and age. Transition rate ratios (TRRs) for LTC accumulation comparing individuals with T2D to those without T2D. TRR was derived from Model 3, which was adjusted for age, time since enrolment, calendar year, sex, current number of LTCs, ethnicity, education, smoking status, and BMI, with interaction terms for T2D × current LTC number and T2D × age, allowing the association of T2D with LTC accumulation to vary by disease burden and age. Point estimates are shown as square markers with 95% confidence intervals represented as shaded bands. *Abbreviations: CI, confidence interval; LTC, long-term conditions; T2D, Type 2 Diabetes; ref, reference*

### Mortality

T2D was associated with higher mortality rates, with the relative excess pronounced at earlier LTC states. Among individuals with 2 LTCs, those with T2D had a 36% higher mortality rate compared to those without T2D (MRR=1.36, 95% CI: 1.28-1.45). Among those with 3 LTCs, T2D was associated with a 23% higher mortality rate (MRR=1.23, 95% CI: 1.17-1.29), and among those with 4 LTCs, an 11% higher mortality rate (MRR=1.11, 95% CI: 1.07-1.15).However, this pattern reversed at higher LTC burdens, with T2D associated with lower mortality rates at 6 LTCs (MRR=0.90, 95%CI:0.88-0.93) and 7+LTCs (MRR=0.82, 95%CI:0.79-0.85). (Table 4).

### Sex stratification

All the models were fitted separately for each sex. Females consistently had higher TRRs than males across all transition states, with the difference being most pronounced in early transitions (1→2: 2.87 vs 2.61; 2→3: 2.41 vs 2.24; 3→4: 2.03 vs 1.92) (Table S4).

### Sensitivity analyses

The results were similar when the number of LTCs were incorporated as factor variables for morbidity progression model and mortality model (Table S5).

## Discussion

In this large population-based cohort study, we found that T2D was consistently associated with accelerated rates of LTC accumulation compared to those without T2D. Notably, this association varied by existing LTC burden and age. The strongest acceleration in LTC accumulation was observed when T2D occurred early in the disease trajectory and at younger ages, indicating that early intervention for T2D may have the greatest potential to prevent or delay subsequent multimorbidity accumulation.

Previous research has primarily examined the association between T2D and the incidence of certain LTC, demonstrating elevated risks for cardiovascular disease (HR∼1.5-3.0)[22], cancer (HR∼1.2-1.5)[23], and dementia (HR∼1.5-2.5)[24]. Beyond individual conditions, cluster analyses have identified T2D as a central node within multimorbidity clusters[2], [5]. At the population level, a Danish study reported that the prevalence of multimorbidity increased from 31.6% to 80.4% over 16 years following diabetes diagnosis[25]. However, these studies are largely limited to a narrow set of preselected conditions, and none has examined how T2D shapes the rate of LTC accumulation spanning a broad range of multimorbidity. To our knowledge, this is the first study to examine T2D in relation to the accumulation of 80 LTCs, allowing us to characterize both the breadth and pace of disease accumulation. We found that T2D was associated with conditions spanning diverse disease categories, including respiratory (e.g., COPD), hematological (e.g.,anemia), and mental health (e.g.,schizophrenia, dementia) among others. These findings, alongside evidence of causal links between T2D and diverse comorbidities[26], suggest that the T2D might act as a key factor of disease accumulation. The diversity of conditions associated with T2D also resonates with the broader epidemiological observation that multimorbidity has shifted over recent decades, driven in part by disproportionate reductions in CVD mortality and a diversification of morbidity burden toward other chronic conditions[4].

A novel feature of our study is the use of multistate models to examine the dynamic role of T2D in LTC accumulation. Unlike traditional approaches that treat comorbidities as fixed baseline covariates, our models incorporate T2D as a time-varying exposure to capture incident T2D throughout follow-up. Critically, by comparing transitions at the same LTC state (e.g., 2→3 LTCs) with and without T2D, we isolated the specific contribution of T2D to disease accumulation while accounting for the total burden of co-existing conditions at each transition point. Furthermore, we tracked disease accumulation across the full spectrum from health to 8+ LTCs, providing a more complete picture of the multimorbidity burden associated with T2D. We found that T2D was consistently associated with accelerated acquisition of additional LTCs across all transition states, with stronger associations at earlier states, suggesting that T2D may be particularly relevant in the early accumulation of multimorbidity. This may reflect the broad pathophysiological influence of T2D, including chronic inflammation, oxidative stress, metabolic dysregulation, and vascular damage, which may lead to the onset of multiple conditions.

Some of this co-occurrence may also be attributable to shared risk factors such as obesity, smoking, or socioeconomic deprivation, which predispose individuals to both T2D and other LTCs[4]. However, a recent bidirectional Mendelian randomization study found that genetic predisposition to T2D causally increases the risk of 23 non-cardiovascular and non-oncologic comorbidities[27], suggesting that the direction of effect runs from T2D towards other conditions rather than being solely explained by shared antecedents. Nevertheless, surveillance bias cannot be fully excluded, as individuals with T2D have more frequent healthcare contact, which may lead to earlier or more frequent diagnosis of additional conditions compared to those without T2D.

We also found that the strongest acceleration occurred in younger individuals at earlier LTC transitions, with progressively smaller association at higher levels of LTC. The attenuation of T2D effects at older ages and advanced LTC states may reflect several mechanisms. Aging-related processes may become the predominant driver of disease accumulation at older age, overshadowing the specific metabolic and vascular effects of T2D. For instance, age-related frailty and physiological function decline affect disease progression regardless of T2D status[28], [29],[30]. Older adults with advanced multimorbidity may develop other severe conditions that become more dominant of morbidity progression and mortality, thereby diminishing the relative contribution of T2D[31]. In contrast, younger people with T2D often present with a different and more aggressive risk factor profile, characterized by more pronounced metabolic dysfunction such as severe obesity, insulin resistance, or fatty liver disease, which could drive more rapid disease progression[32]. Lastly, survivor bias may play a role, individuals with T2D who survive to older ages and accumulate many conditions may represent a selected group with greater resilience or better disease management.

Sex-stratified analyses revealed a stronger relative association of T2D in women than men. This finding is consistent with evidence that women with T2D carry a disproportionately higher comorbidity burden than their male counterparts[33], [34]. A large Austrian study of over 9 million individuals followed for 17 years found that women faced a disproportionately higher risk of progressing to cardiovascular disease once diagnosed with diabetes, hypertension, or metabolic disorders [35]. This sex difference may reflect biological vulnerabilities, including hormonal changes after menopause, sex-specific inflammatory responses, and differences in body composition, as well as differential healthcare-seeking behaviors and management[36].

### Strengths and limitations

A key strength of this study is the novel application of multistate models to examine the dynamic role of T2D in LTC accumulation. By modelling transitions across successive disease states and accounting for the competing risk of death, this approach enabled us to decompose the transition process around T2D diagnosis and estimate its dynamic effect on disease accumulation. The results demonstrate that T2D consistently accelerates LTC accumulation regardless of the disease state at which it occurs. This multistate approach moves beyond traditional Cox regression models, capturing the dynamic and interdependent nature of disease transitions. Notably, the framework holds broader potential and can in principle be extended to any chronic condition of interest, offering a versatile analytical template for future research into the dynamics of multimorbidity. In addition, we considered a broad spectrum of 80 LTCs, including both concordant diabetes-related complications and disconcordant conditions[5], providing a more comprehensive characterization of multimorbidity in T2D. The study also benefited from a large sample size and extended follow-up period, which provided sufficient statistical power to reliably estimate transition rates across all LTC accumulation states.

There are several limitations of our study. First, we treated each LTC as equally contributing to MLTC progression, which may not reflect clinical reality where some conditions are more severe or impactful than others. Future research could consider a weighted approach that accounts for disease severity, healthcare utilization intensity, or prognostic impact when examining multimorbidity progression. Secondly, the disease diagnosis information was identified via ICD code using health electronic records, which might underestimate certain conditions, or only capture the severe cases[37]. Similarly, detection bias may influence the results, as individuals with T2D receive more frequent medical monitoring, potentially leading to earlier identification of additional chronic conditions[38]. However, as our results are presented on the relative scale, any potential bias is likely to be less differential We were also unable to examine the mechanisms underlying the accelerated multimorbidity accumulation observed in individuals with T2D. Factors such as glycemic control, diabetes duration, and age at diagnosis may modify disease progression rates and warrant investigation in future studies. Finally, participants in the UK Biobank are generally more well-educated and healthier than the general population, thus our findings may not be generalizable to other populations with different sociodemographic characteristics [39].

## Conclusion

Our study suggests that T2D is associated with accelerated development of subsequent multimorbidity, highlighting T2D as a critical contributor of chronic disease accumulation. The association was stronger at early multimorbidity state and younger age, suggesting that early prevention and management of T2D may be an important strategy in delaying the cascade of multimorbidity development. However, the mechanisms underlying the stronger associations observed at earlier multimorbidity states require further investigation.

## Data availability

UK Biobank data are available to researchers through the standard application process (https://www.ukbiobank.ac.uk/enable-your-research/apply-for-access). This study was conducted under UK Biobank Application Number 81520.

## Abbreviations

BMI: body mass index
CHD: Coronary Heart Disease
COPD: Chronic Obstructive Pulmonary Disease
HR: hazard ratio
IQR: interquartile range
LTC: long-term conditions
MLTC: multiple long-term conditions
MRR: mortality rate ratio
PY: person-years
SD: standard deviation
STROBE: 
T2D: type 2 diabetes
TRR: transition rate ratio
UKB: UK Biobank

## Human Ethics

The UK Biobank study received ethical approval from the North West Multi-centre Research Ethics Committee (MREC reference: 11/NW/0382). All participants gave written informed consent before enrolment in the study, which was conducted in accordance with the principles of the Declaration of Helsinki. This study has been conducted using the UK Biobank Resource under Application Number 81520.

## Acknowledgement

We thank the Steno Diabetes Center Aarhus for providing financial support to access UK Biobank data. The authors acknowledge the contributions of all study participants and the research. We also thank Luke Johnston for developing the UKBAid R package, which was instrumental in our data analysis process.

## Contribution statement

JZ: Conceptualisation, formal Analysis, and original draft.

LB, DW: Conceptualisation.

BC, DW: Supervision of the analytical process.

LB, SBG, HS, BC, DW: Interpretation of results.

All authors reviewed and edited the manuscript and approved the final version for submission

---

**Figure S1.**
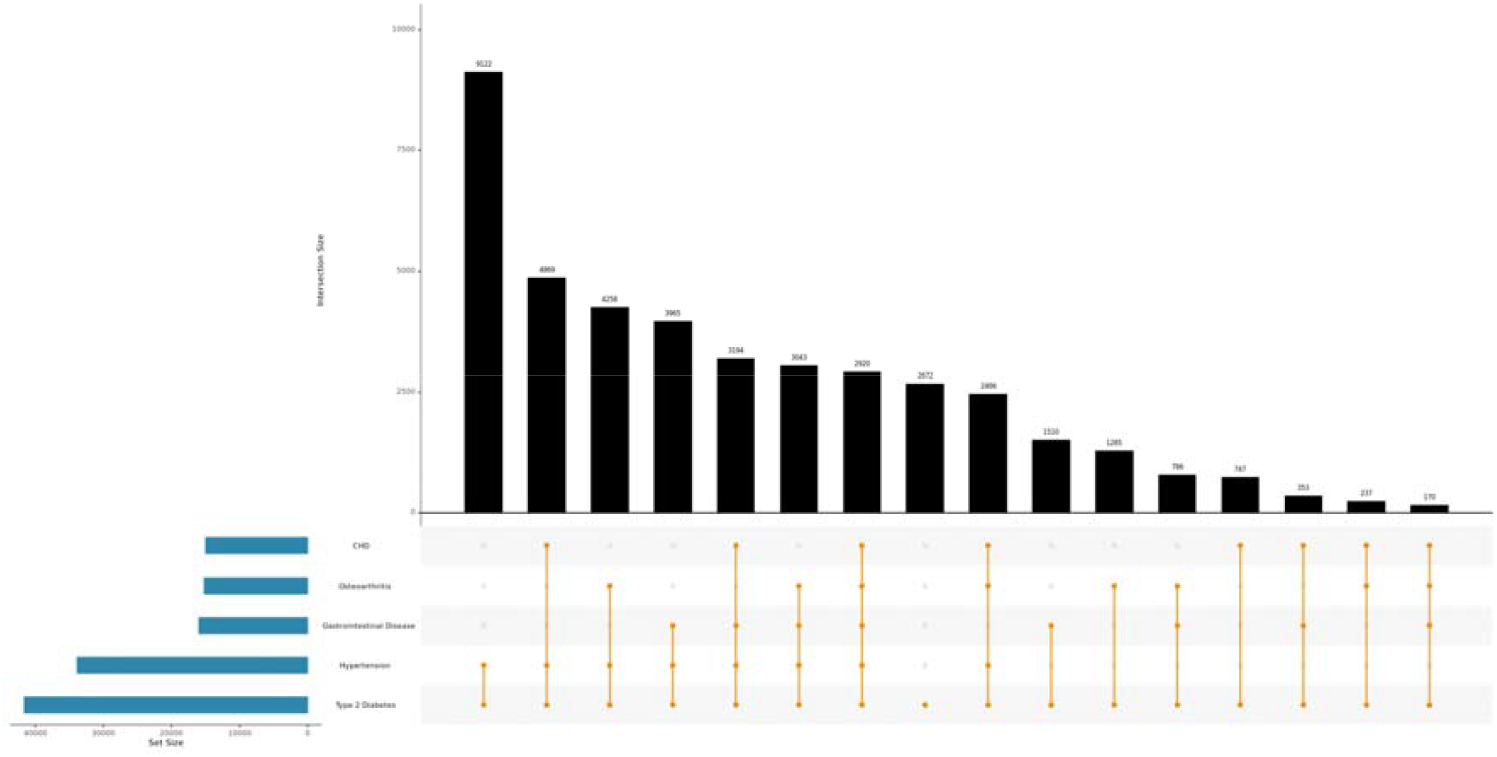
Cumulative co-occurrence of T2D with other LTCs

